# Migration and the Emergence of Chagas Disease Vectors in the Growing City of El Pedregal, Peru

**DOI:** 10.1101/2023.08.08.23293548

**Authors:** Raquel Gonçalves, Kathryn P. Hacker, Carlos Condori, Sherrie Xie, Katty Borrini-Mayori, Lina Mollesaca Riveros, Roger Quispe Apaza, Manuel Ysidro Arratea, Gustavo Nativio, Ricardo Castillo-Neyra, Valerie A. Paz-Soldan, Michael Z. Levy

**Author notes:** Corresponding author: Michael Z. Levy.

## Abstract

The city of El Pedregal grew out of a desert, following an agricultural irrigation project in Southern Peru. We conducted door-to-door entomological surveys to document the emergence of triatomines and bed bugs into this new urban environment. We inspected 5,191 households for *Triatoma infestans* (known locally as the *Chirimacha*); 21 (0.41%) were infested. These were extremely spatially clustered (Ripley’s K p-value <0.001 at various spatial scales). Using remote sensing we compared the year of construction of infested to un-infested households and found that infested houses were older than controls (Wilcoxon rank-sum: W=33; p=0.02). We confirmed infestations through a subsequent bed bug specific inspection in 34 households. These households were more spatially disperse across El Pedregal. To gain a better understanding of the context surrounding triatomine infestations, we conducted in-depth interviews with residents to explore their migration histories and previous experiences with *Chirimachas*. Main reasons for migration includes searching for work on land, opportunity to buying a house, and scape adverse climate effects. Permanent migration flow and poor housing conditions create suitable environment for emergence triatomine infestation. We discuss how changes in the landscape could potentially heighten vulnerability to vector-borne illnesses.

**Author summary:** Large-scale irrigation and changes in land-use have been linked with emergence of infectious disease worldwide. In El Pedregal, Southern Peru, the Majes-Siguas irrigation was designed to supply water to agribusiness companies installed in a desert area. This project has propelled a constant migration flow and the growth of this new city, promoting conditions to emergence of *Triatoma infestans*, an insect vector of Chagas disease, as well as bed bugs. Triatomine infestation presented a clustered pattern, its dispersion limited by unoccupied houses. Triatomine infested houses tend to be older than other houses. Bed bug infestations were more spread out, and not related to construction age. Householders’ stories of migration shed light on some of the socioeconomic determinants that promote conditions for infestation. Amongst these, living in poor housing conditions, and the constant migration flow; driven by seeking opportunities to work, to buy a house, or to escape from adverse climate effects in other farming regions. Environmental and socioeconomic impacts of large-scale development disproportionately affect vulnerable populations. Design of surveillance systems that account for these contexts is urgently needed to support early detection and control of emergence of vector-borne diseases.

## Introduction

Some great cities of the world have mythical origin stories; the origins of others are more mundane. In many cases the impetus for a *de novo* city is political—national capitals like Washington DC (1) and Brasilia (2) were placed in central locations, and the blank slates allowed for national architectural expression (1). In many other cases, cities grow *de novo* in response to the exploitation or cultivation of new economic resources, such as rubber (Manaus, Belem, Iquitos) (3), gold (San Francisco) (4), oil (Houston, Texas USA) (5), among many others. The rapid emergence of entirely new cities allows for the close study of the processes of arrival and spread of urban pests, including disease vectors.

The foundation of new cities increases human migration, mainly as a workforce, and can act as a persistent social determinant in the processes of introduction and emergence of vector-borne diseases (6,7). Further landscape changes, such as expansion of agricultural frontiers, and changes in land use, provide broad opportunities for new contacts among hosts, vectors, and pathogens (8). Socioeconomic factors and the poorly planned occupation of spaces associated with poverty, are structural processes that can promote conditions for infestation by triatomine bugs, the vectors of the parasite *Trypanosoma cruzi*, causative agent of Chagas disease. Poor housing conditions, such as poor quality construction materials, cracks on the walls, unplastered surfaces, and presence of animals are factors associated with increased risk of triatomine infestation (9,10).

In southern Peru, a new city, El Pedregal, was created *de novo* following an irrigation project that created numerous job opportunities attracting migrants from local towns as well as more distant regions. We studied the early stages of emergence of the insect vector of Chagas disease, *Triatoma infestans*, in El Pedregal as well as bed bugs, *Cimex* sp., which are also competent vectors of *Trypanosoma cruzi* (11–13). We performed door-to-door entomological surveys throughout El Pedregal, and, in concert with targeted insecticide application, conducted in-depth interviews with affected residents and their neighbors to examine the economic and social forces that led to the rapid growth of the city in a barren desert.

### Study site

The study was conducted in Villa El Pedregal (−16.32699, −72.19480), Distrito de Majes, Provincia de Caylloma, Departamento de Arequipa, Perú. Located in the Subtropical Coastal desert (14) the city sits at an altitude of around 500m, and boasts a year-round temperate climate (mean annual temperature of 19°C), but with very little precipitation (150mm annual mean) (15). In 2017, the Peruvian census for Majes district reported 60,108 inhabitants.

In the 70’s, the Peruvian Government initiated an irrigation project, ’*Proyecto Especial Majes-Siguas*’, with the goal of providing a water source for the irrigation of 60,000 hectares destined to agroindustry in the ’Pampas de Majes’. The first phase of the project consisted of the construction of the Condoroma dam, and channelling (and in some cases tunnelling) water from the the Colca River to the Siguas River, and from there to Pampas de Majes. By the 1980s the project had supported the establishment of agroindustry and processing plants in El Pedregal. These in turn attracted subsistence farmers from different parts of Peru, who came to the region seeking work opportunities (16).

## Methods

### Entomological surveys

In collaboration with the Ministry of Health of Arequipa we conducted at-the-door interviews and entomological inspections in El Pedregal between October, 2019 and April, 2022, with numerous disruptions due to the Covid-19 pandemic. These surveys were meant to assess the distribution of triatomine vectors for the purposes of planning indoor residual spraying (IRS) to eliminate them. We asked several questions at the door regarding individuals’ experience with triatomines and bed bugs. We then requested permission to perform a full inspection of the house, both inside and outside, for triatomines. We separately offered to inspect beds and furniture for bed bugs to those individuals who reported bites or suspected an infestation. Prior to the entomological surveys, we created a georeferenced map of all households and other structures and integrated entomological data into this map in real time using the Vectorpoint app – an app created by our research team for use by house inspectors to guide entomological searches (17). We informed the Ministry of Health of all households in which we detected triatomines, and, in coordination with them, scheduled the households and their immediate neighbors for insecticide treatment per Ministry of Health guidelines.

### Indoor Residual Spray (IRS)

IRS was offered to all householders where triatomine infestation was confirmed. In the blocks that infestation was confirmed in more than one household, IRS was offered to all houses within block, and to households in the neighboring blocks that faced the treated block.

IRS was conducted twice per household at a target 6-month interval, though the pandemic led to a longer delay between applications. The first phase (IRS-1) was conducted between November 20th, 2020 and March 17th, 2021; 135 households were treated. The second phase (IRS-2) was conducted between October 5th, 2021 and February 13th, 2022, and encompassed 118 households. All participating households were treated with Lambda-cyhalothrin SC 10% (Arpon® 10 SC) in a dilution of 60ml of insecticide: 8 Litres of water, according to the manufacture’s indications. The insecticide was provided by the Ministry of Health. Vector control technicians applied the insecticide using a Hudson® sprayer tank (H.D. Hudson Manufacturing Company, Chicago, IL), with 8.5L capacity (Pressure 60 P.S.I.; 414 kPA) (Fig. 1).

**Figure 1:**
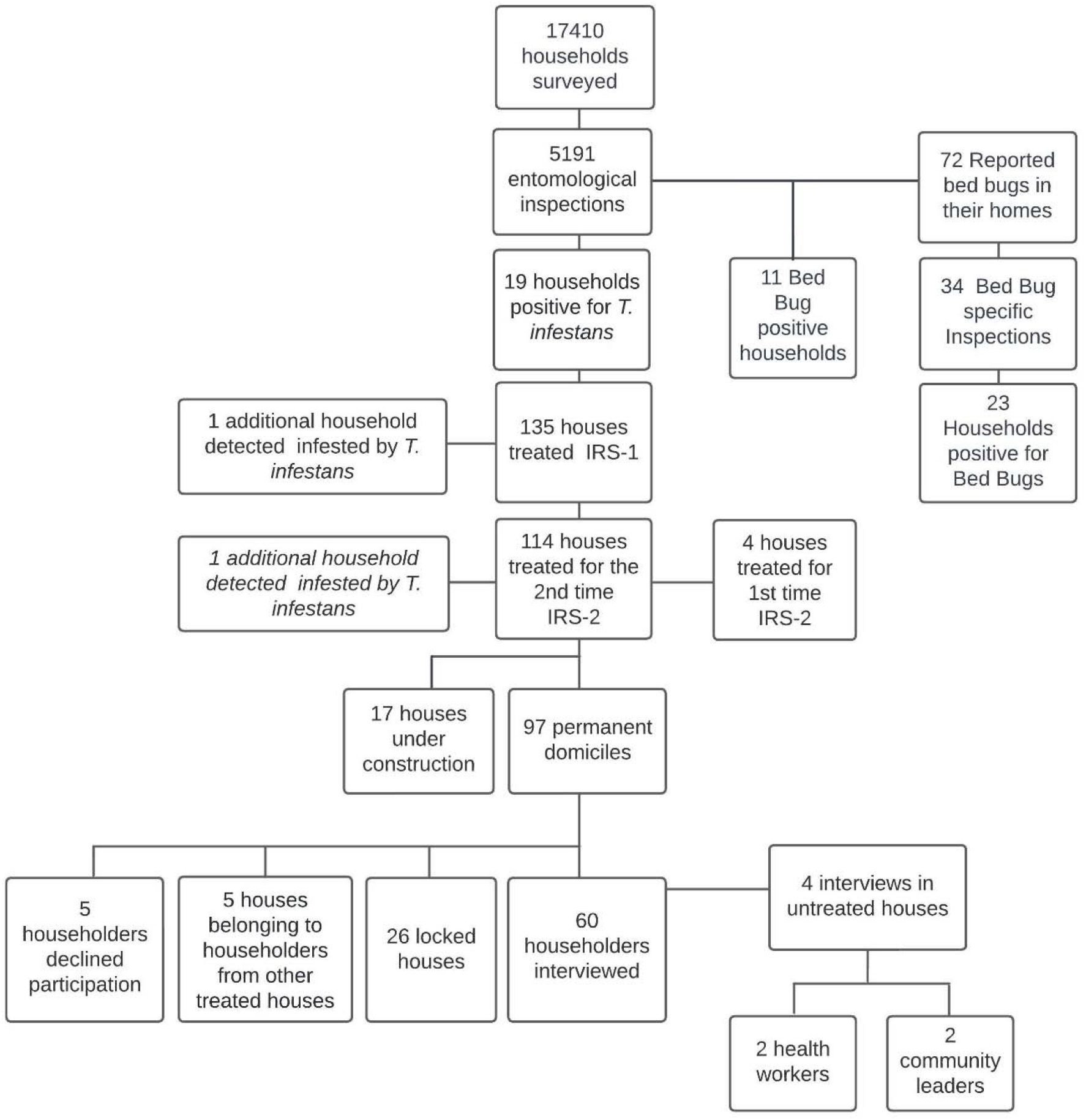
Flowchart of entomological acti vities and in-depth interviews conducted in El Pedregal.

### Semi-structured interviews

We visited 97 houses (of 118) treated during the 2nd IRS application. Houses were visited up to three times, in different work hours and during weekends. A total of 65 householders were contacted, and individuals 18 years or older were invited to participate in semi-structured interviews. Sixty individuals agreed to participate, while 5 declined participation. Additionally, two community health workers and 2 community leaders were invited to participate to provide additional context surrounding vector infestation and control in the region (Fig. 1).

A semi-structured interview guide was developed to ensure that all interviews covered similar topics and was organized in three main areas of interest: 1) migration story, 2) resident perceptions regarding the city’s growth, and 3) experiences and knowledge related to dangerous insects, specifically focusing on triatomines and bed bugs. All interviews were recorded; one of us (RG) listened to each at least twice to add detail to handwritten field notes. A matrix summarising themes that emerged inductively associated to the three main areas of interest was created manually; this matrix was used to generate the summary of themes and quantify how often these topics emerged. For additional detail and sample quotes, thirty-five audios were transcribed (n=25 randomly selected and an additional n=10 selected by the authors due to their richness), uploaded to Dedoose (18), and coded. Coding was both deductive (i.e., based on the main themes explored) and inductive (based on responses that emerged from interviews). Themes and related quotes are summarised and presented in the results section.

### Assessment of Household Development via Satellite Images

A sample of 142 uninfested houses (2.75% of total surveyed houses n=5,164) was randomly selected in order to compare the median year of construction with houses infested by *T. infestans,* and with houses infested by *Cimex* sp. The estimated statistical power to detect difference between medians was 80% (alpha=0.05) by application of non-parametric Wilcoxon rank-sum test. The set of houses was assessed in Google Earth images, obtained for the years 2004, 2010, 2012, 2017, 2018, and 2020. A matrix was generated, registering for each house the year in which images indicated first evidence of development, defined as the presence of a building covering any portion of the sample city lot.

### Spatial Analysis

We assessed spatial clustering of triatomine and bed bug infestations using Ripley’s K function, which compares a given point distribution to complete spatial randomness to determine if points are clustered, dispersed, or occur at random (19). Separate K functions were used to examine point patterns of cases (inspected houses that were positive for the infestation of interest) and controls (inspected houses that were negative for the infestation of interest) for both triatomines and bed bugs for a distance of 0 to 1000 metres. For each infestation type, we calculated the difference between K functions (KD) for cases and controls to assess for clustering of cases while accounting for underlying spatial heterogeneity in the locations of inspected houses; positive KD values indicate significant clustering of cases. Monte Carlo random labelling simulations (n = 999) were performed to determine 95% confidence intervals to assess statistical significance for alpha level = 0.05. Spatial analyses were conducted in R using the spatstat and smacpod libraries (20,21).

### Ethical approval

The study was approved by the ethical review board of the Universidad Peruana Cayetano Heredia (protocol #103096), the University of Pennsylvania (protocol #833122), and the Tulane School of Public Health and Tropical Medicine (#2019-909). Individuals who agreed to participate in the semi-structured interviews signed a consent form, authorising recording of the interviews. Entomological surveys and insecticide application conducted as part of the Ministry of Health’s control campaign were deemed not human subjects research and were conducted following verbal consent in accord with the Ministry of Health’s usual protocols.

## Results

### Entomological surveys

We conducted visits to 17,410 households to survey for triatomine bugs. We were able to interview household members at the door of 12,129 houses (69.7%) and enter and inspect 5,191 (29.81%). We confirmed infestation by *T. infestans* in 19. All households infested by *T. infestans*, as well as their immediate neighbors, were requested to allow indoor residual insecticide (IRS) treatment. During these treatments we detected triatomines in an additional 2 households, bringing the total number of infested households to 21 (0.41% of those inspected).

During the at-the-door interviews we asked household members about potential infestations with bed bugs. We presented a Petri dish with bed bug specimens at different development phases. We found that 619 householders recognized bed bugs, 72 reported that they had seen a bed bug in their house, and 53 reported that they had been bitten by bed bugs in the last year. Of the 53 who reported recent exposure to bed bugs 34 requested bed bug specific inspections (including beds, furniture, and other likely harbourages). We detected bed bugs in 23 of these houses.

Infested triatomine households were extremely spatially clustered; the 21 total infestations were located on only 6 city blocks. Four of these blocks were themselves clustered together in two neighborhoods, and one household was not obviously associated with either focus of infestation. By contrast households infested with bed bugs were more widely dispersed, though still clustered in some areas.

A total of 258 triatomines were collected in the 21 positive households [median: 5 per household, IQR: 2, 14.5]. Eleven households had insects solely in peri-domestic spaces; 5 had insects solely inside in domestic spaces, and 5 had both. Nymphs were captured in 71% of infested houses indicating colonisation by the vector. We microscopically examined a total of 215 of the 258 (83.3%) specimens for *Try. cruzi* infection (the remaining insects were too damaged or too dry to examine) and all were negative. The lack of *Try. cruzi* in these insects is in line with our observations in the city of Arequipa, where we have not had parasite-infected insects in areas with very sparse vector infestation (22,23).

### Semi-structured interviews

#### Socio-demographic and migration patterns

Table 1 presents participants demographic information, work at arrival in El Pedregal, and migration history. The majority of migrants (53.1%) reported repeated migration movements, living in at least one city different from their origin place before their move to El Pedregal.

**Table 1.** Participants demographic, migration history and work at arrival in El Pedregal.

| ***Participants*** | **Median age (IQR)** |  |
| --- | --- | --- |
| Female (n=44) | 39.5 (30.2; 48) |  |
| Male (n=20) | 40 (27; 58) |  |
| ***Place of origin (Department level)*** | **n (%)** |  |
| Apurimac | 1 (1.56) |  |
| Arequipa | 25 (39.06) |  |
| Arequipa (born in El Pedregal) | 5 (7.81) |  |
| Ayacucho | 1 (1.56) |  |
| Cusco | 12 (18.75) |  |
| Moquegua | 1 (1.56) |  |
| Huánuco | 1 (1.56) |  |
| Puno | 18 (28.13) |  |
| ***Education level*** | **Female n (%)** | **Male n (%)** |
| Uncompleted primary | 4 (9.1) | 2 (10) |
| Completed primary | 3 (6.8) | 2 (10) |
| Uncompleted secondary | 3 (6.8) | 3 (15) |
| Completed secondary | 11 (25) | 5 (25) |
| Institute/technical education | 3 (6.8) | 1 (5) |
| Graduated | 4 (9.1) | 3 (15) |
| Missing information | 16 (36.4) | 4 (20) |
| ***Work at arrival in El Pedregal*** | **Female n (%)** | **Male n (%)** |
| Agriculture | 12 (27.3) | 8 (40) |
| Agriculture and commerce | 3 (6.8) | 0 |
| Agribusiness companies | 4 (9.1) | 3 (15) |
| Commerce and services | 8 (18.2) | 3 (15) |
| Housewives | 8 (18.2) | 0 |
| Arrived as child | 6 (13.6) | 3 (15) |
| Born in El Pedregal | 3 (6.8) | 2 (10) |
| Missing information | 0 | 1 (5) |
| ***Migration history*** | **n (%)** |  |
| Born in El Pedregal and has never left | 5 (7.8) |  |
| Migrated from their place of origin straight to El Pedregal | 21 (32.8) |  |
| Migrated through more than 1 local apart from their place of origin before moving to El Pedregal | 34 (53.1) |  |
| Migrated from their place of origin to El Pedregal, left, and returned to El Pedregal (back-forth) | 4 (6.3) |  |

#### Work at arrival: the agribusiness companies and changes in agricultural practices

Upon arrival to El Pedregal, about half of the migrants interviewed reported working in agriculture (Table 1), either in “parcelas” (i.e., a portion of agriculture terrain of 5 hectares that are part of a bigger farming unit) or in the crop companies.

Initially, in the 1980’s, agricultural activities were focused around raising cattle for milk production. The first settlers received credits and imported animals to support this production. Participants also recounted how one of the first agribusiness companies established in El Pedregal controlled the milk price and, together with the high prices of supplies needed for this activity, made it unsustainable for small farmers to compete. Participants recalled having to sell their cattle and dedicate themselves to other agricultural activities.

> *“… With dairy cows, the colonists with dairy cows, there was very little agriculture. It was a rare few that grew carrots or onions. There was no other farming… The majority of people worked with dairy cows because there was a credit system that facilitated the colonists to obtain cows, imported cows.”*

> *“The people dedicated themselves to dairy because this was a dairy zone; cattle, to the sale of milk, the fattening of cattle, was the principal source of income here in the Majes irrigation. Unfortunately, with the price the company… that had the monopoly, of course [they] always had been here, and the elevated cost of supplies, it was a losing battle. Many gave up on cattle, it’s no longer sustainable to keep up that business.”*

> “…*I had cattle and I thought ’What do I do with my cattle? Because they don’t bring in money…Most of my friends that had cattle gave up on the cows”*

#### Reasons for migration to El Pedregal

Migrants perceived – and moved to — El Pedregal as a place with opportunities to improve their lives; with more permanent opportunities to work, to build their homes, and to provide education for their children. Some migrated to El Pedregal to reunite with a family member already established there; others migrated from locations with seasonal work activities, such as those from Puno who mentioned that agriculture and cattle raising was seasonal, compared to opportunities in El Pedregal where the cultivation of different plant species provided year-round employment.

> *“… Because where I am from (Puno), there is not much work. Here in Pedregal there are jobs, of all kinds… There [in Puno] I only dedicated myself to my livestock. I would watch my livestock because I did not have much work. Livestock is sold annually, not monthly… I also have 5 children and I have come here to be able to support my children, to have them study.”*

The demand for goods increased as the population increased, boosting the local economy, and generating additional work opportunities in commerce, construction, and various service industries. The stability of these permanent year-round labor opportunities combined with the possibility to participate in “loteamiento” (procurement of land lots) in El Pedregal, were reported as reasons for migration.

Climatic conditions were also mentioned as a reason for migration. In Puno, for example, climate events such as flooding affect crops, lives, and food security in the recent years (24); the climate in El Pedregal is considered appropriate year-round for different agricultural activities.

> *“… Over there [Puno] we suffered from flooding… floods have taken all the crops, that is why we have come, we arrived looking for work… Now everything is being flooded over there…”*

> *“Firstly, this is an irrigation, an irrigation where everything grows. The people count on this. The climate is appropriate for everything, raising animals… in 40 days the chickens are ready. For this they come, the climate is very good… The production grows quickly, you have three crops a year.”*

#### Changes in landscape, land ownership, and housing

Participants reported that when the irrigation project started, El Pedregal was a desert. There were no houses, and all the people used to live in the *parcelas.* The *parcela* owners (locally called “settlers” or ’colonists’), hired the migrant farmers to live and work on their lands. The first populated area in El Pedregal was the La Colina neighborhood (one of the tree neighborhoods where triatomine infestation was detected), and the first constructions there were dedicated as “sedes” (headquarters) for a cooperative, the agrarian bank, and to a milk company, which used to buy milk from the farmers and, through their headquarters in La Colina, to make the respective payments. Over time, people applied for formal tenancy lots (land title or similar recognition). The time frame between land acquisition and tenancy is reported to be long in the newest settled areas, particularly due to the lack of infrastructure. In order to push for permanent tenancy, land acquisition was sometimes conditioned on its immediate occupation. As arriving migrants had few resources, they constructed shelters for themselves with fragile materials (e.g. “*esteras*” which are woven straw mats), but as they gradually become settled, with titles of land ownership or access to electricity, they invested in materials for more permanent house construction, which generates job opportunities for those in construction. First local schools, health facilities, and food markets were also recalled being constructed with *esteras*, and rocks, replaced by plywood.

The absence of permanent neighbors is recalled in the past, and still reported in the present in newly settled areas (where triatomine infestation was also detected), where at the time of interviews, there is still no public water supply.

> *“… we bought the lot and they told us ’you have to live on the lot because if not we’ll take it’ … So all the houses were just 4 esteras [Woven reed mats], there was nothing around the lot. Now people with a little money close their lot in”*

> *“At first we bought esteras [Woven reed mats] … it was hard, I slept on a doubled-over estera, we folded an estera and my children slept below … It has been hard… like everyone we started like that…”*

> *“… Not even the market was like that, it was just some fabrics, the health post also was made of esteras, it wasn’t even concrete, just rustic, later they made it of wood.”*

> *“… Modulo G was empty land, not a single house. Ours was a house of little esteras, with a little room… the school wasn’t there, later bit by bit [the urbanization] formed, the rooms were of plywood and later they constructed them. Still people didn’t live here, we didn’t have any neighbors, we still don’t…”*

#### Occurrence of dangerous insects and experiences regarding triatomine and bed bug infestation

When asked about occurrences of dangerous insects in their community, participants recalled seeing triatomines at their homes among other insects and arachnids (e.g., spiders, scorpions, flies, mosquitoes, ticks). The oldest inhabitants reported triatomine infestation in different neighborhoods of El Pedregal (e.g., Pedregal Norte, Alto, Colina, Malvinas, and in the *parcelas*, particularly in the animal paddocks) and in the neighboring locality of Santa Rita de Siguas in the late 1980’s/early 1990’s. Frequent insecticide spraying campaigns are recalled having controlled triatomine infestations.

Migrants demonstrated knowledge regarding risk factors for triatomine infestation, particularly related to poor housing conditions and breeding animals near human dwellings. Participants knew they had to report infestation at the health centre facility, and posters and flyers displaying pictures of triatomines were mentioned as sources of information about how to identify the insect and how to report infestation.

> *“… I’ll tell you when, in 1986, 1987, there were tons. I lived in a parcel, I worked in a parcel, and when I arrived on the parcel itself there were tons and here in Pedregal the people complained a lot about Chirimachas. There was a fumigation company, they fumigated frequently, after that the bugs disappeared.”*

> *“I knew of the Chirimacha because, when we lived in the valley, we used to see lots In the valley [The Valley of Siguas] there are strong campaigns for Chagas disease, because there are tons of Chirimachas. The majority of the houses were of adobe and in the adobe lived the Chirimachas. The thing is, in the valley we had all our houses tied to the hillside, and well, the hills were of that red earth and there always were Chirimachas. Although in our house there weren’t many because, as I raised roosters, I always fumigated. But one year they went to the neighbor who raised guinea pigs on the roof of her house. She made her house out of cement, supposedly more hygienic than my house that was of adobe, and then they fumigated her house in her guinea pig pens and waaaa how they came down! It looked like the procession of the Señor de los Milagros*, tons. We never saw so many Chirimachas as that day they fumigated the house of our neighbor.”*

> *\*the procession of the Señor de los Milagros is a religious procession that occurs in Peru each October*

> *“It was a big black thing, with a kind of swollen belly, I looked at it, I had never seen it before. I saw it on the wall when we turned on the light… What could it be? I didn’t know, what animal could it be As I often went with my daughter to the health post, I went there and I had seen a sign that said ’Chirimacha’ and that it produced a disease, Chaga. So I left it there.”*

> *” I came to know it here because where I slept was made of esteras, while I slept, they walked all over me, on my face, and I’d turn on the light and see the Chirimachas. “*

Participants reported having heard about bed bug infestations from their neighbors, friends, and health workers in door-to door prevention campaigns. Inhabitants who had their houses infested by bed bugs, reported use of different products and practices to control infestation, such as: use of “Sapolio® aerosol”, “Bolfo®” (Propoxur-carbamates), chlorine, and cleaning activities (taking out the mattress to expose it to the sun). Some of those who experienced infestation believe that bedbugs may transmit diseases but did not know which one.

> *“…I moved recently, a year ago, and bumps appeared… I washed, cleaned and there was nothing, you can’t see them “G-d what is this? and a friend told me ’the bed bugs bite you that way in one spot, lots in one spot’. She asked me to show her the bumps and she told me that it was bed bugs, I was frightened, I asked her ’What do I have to do? Then she told me about a sign on a kiosk that said, ’Call when you have Chirimahcas’. I called then and I told them ’Please, I don’t know what I have’ so they came and inspected everything, the women took apart my bed and there they were, inside. I had to use bleach, boiling water; I didn’t want to sleep there anymore”.*

> *“…Because every little animal passes from neighbor to neighbor…If they had here those bugs I’d use Bolfo®, It doesn’t cause harm, and put everything out in the sun, more than anything, put things in the sun, like for fleas, there used to be tons of fleas, we had to put the blankets out in the son, and as the sun warmed the covers the fleas walked off, they left to search for a place to hide themselves… because at night you can’t see them… But if you spread the covers out in the sun they leave. They might come from the mountains in the things that travellers bring, they could come in clothing, in the bedding, in anything that they bring they could come and I understand that this little animal adapts itself to any environment, from cold to hot…”*

Although participants had their houses IRS treated, 79.3% (23/29) reported having used other insecticidal products, such as: Butox® (deltamethrin) against spiders and mosquitoes; Raid® against chicken ectoparasites; Sapolio® against spiders, mosquitoes, flies, triatomines and bed bugs; Baygon® against spiders and flies. Two respondents reported using the agricultural pesticide Lannate® (Methomyl-carbamate), applied indoors and outdoors (against triatomine infestations), and applied directly on a dog (for fleas).

> *“… if there were a lot of [triatomines] we definitely have to fumigate…What we used was what we used on the fields, those sprayers the work by pressure, spraying with those, relying on a special product for Chirimachas, I dont remember which. There are various products that have various names, we used ’Lannate [Methomyl], which was a strong poison, we sprayed with that and we had to be outside the house for at least 12 hours.”*

> *“In La Joya we sprayed, but I can’t remember the name [of the insecticide]. We haven’t sprayed here… A long time ago I had a dog… It got fleas and couldn’t get rid of them…I tried everything until a neighbor told me to use Lannate, which they use on the farms, and then I could kill the fleas.”*

#### Assessment of Household Development via Satellite Images

Houses infested by *T. infestans* were constructed significantly earlier than control houses (Wilcoxon rank-sum test: W=33, p=0.02). No statistical difference was observed between median years of construction when we compared houses infested by *Cimex* sp. and control houses (W=6.5, p=0.07).

#### Spatial Analysis

The difference in K functions (KD) indicated significant clustering of both triatomine and bed bug infestations at various spatial scales (Fig. 2). The Ripley’s K functions of observed case and control data for both infestation types showed significant clustering across all distances considered (Fig. 2A-2B and 2D-2E), suggesting that the locations of inspected houses exhibit significant clustering at baseline. However, the observed KD functions were positive up to 1000m for triatomines and up to 600m for bed bugs, which suggests that triatomine and bed bug infestation exhibit clustering over and above any clustering observed in non-infested homes (Fig. 2C and 2F).

**Figure 2.**
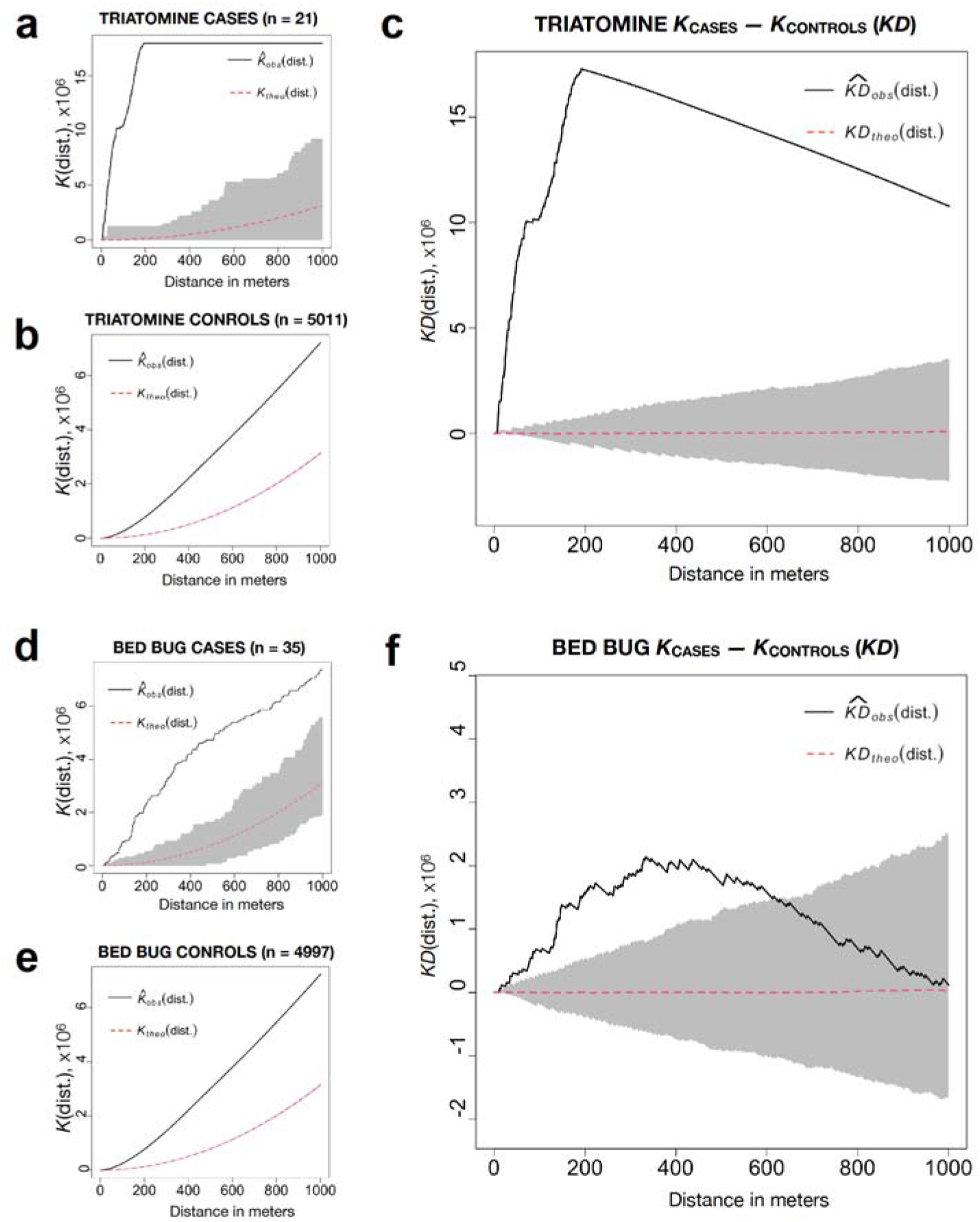
Ripley’s K functions measuring spatial clustering for triatomine cases (a) and controls (b) and bed bug cases (d) and controls (e), as well as the difference in K functions (KD) for cases and controls for triatomines (c) and bed bugs (f) for a distance of 0-1000m. Black lines indicate observed values, dotted red lines represent null hypothesis of no clustering, and grey shading represent 95% tolerance envelopes. Significant clustering for triatomine infestations is detected at all distances, while clustering for bed bug infestations is detected for distances less than ∼600m.

## Discussion

Throughout history, human populations have employed migration as a survival strategy in response to various challenges, including the impacts of climatic events (26–28), food insecurity (29,30), or pursuing improved living conditions (31). Governmental policies focused on the implementation of extensive infrastructure projects, extractive industries, and agribusiness activities, can generate a need for additional labor force (32), often leading to migration into newly urbanizing areas. The resulting, often haphazard, settlement promotes conditions conducive to disease transmission by connecting hosts, pathogens, and vectors (33).

The *Chirimacha*, *Triatoma infestans,* successfully invaded urban and peri-urban areas in Arequipa, Peru (10,23); Cochabamba, Bolivia (34); São Paulo, Brazil (35); and Argentina (36,37); complicating regional efforts to eliminate vector-borne Chagas disease (23,37). Here, we describe the migration stories that contributed to population growth in El Pedregal, and we assess residents’ perceptions regarding urban development, and their experiences regarding triatomine and bed bug infestations.

The Majes irrigation project was designed to supply water resources for the establishment of agribusiness companies in El Pedregal (16). These companies had a huge impact in local farming practices and economy, by introducing new products and agriculture practices, fixing prices for milk, and eventually leading to a shift in the main products in this region: from cattle and dairy to fruits and vegetables. Their establishment required a permanent labor force, which attracted subsistence farmers from across the country. These populations declared different reasons for migrating to El Pedregal; while some of them were seeking opportunities to own a house or to improve their living conditions, others were only searching for a new place to keeping working the land. Subsistence farmers assumed the agribusiness companies would secure a permanent work offer, a different reality than they faced elsewhere, where their farming practices were vulnerable to seasonality, local trade dynamics, and to adverse climate events. Coming to a desert, these migrants arrived with few resources; only enough to live in shelters constructed of woven reeds and other poor-quality materials. Over time, working in the fields with a fairly consistent income, they constructed more permanent houses. Most of the houses, however, continued to incorporate precarious materials, such as unmortared stone. The prolonged precarity of houses in the area provided stable environments for introduced triatomines to establish colonies.

Through interviews and satellite images, we assessed house construction and the expansion of the settlement over space and time. We observed that not all constructed houses were immediately inhabited, as evidenced by the nearly one-to-one inhabitant-to-dwelling ratio in the 2017 Peruvian census for Majes district, which reported 60,108 inhabitants and 58,298 dwellings registered (38). This low ratio is likely due to the large number of dwellings that have been ’staked out’ but not fully constructed. In our door-to-door surveys in the fairly established areas, we found that approximately 15% of houses were uninhabited. The long interval between the tenancy of space and its effective use was also mentioned by participants in the in-depth interviews. Uninhabited neighboring houses might act as a limiting factor for triatomine dispersion, as few sources of food are available in these structures (39). The uneven construction may thereby have restricted triatomines to a small number of households, spatially clustered, in the older sections of El Pedregal.

Households infested with *T. infestans* tended to be older, in areas where migrants had come many years ago. In contrast, infestation by bed bugs presented a different pattern, more dispersed in comparison to those infested by *T. infestans*. Bed bug infestation was not associated with the age of the home, nor the level of construction of the structure. Although bed bug infested households were more spatially clustered than would occur by chance, they were much more widely dispersed across El Pedregal than the triatomines. Bed bugs are likely less dependent on active dispersal in El Pedregal, and, by being carried passively in belongings, less affected by the barriers of uninhabited houses, barren vacant lots, and streets. Awareness of bed bug infestation was amplified through communication amongst community members and health workers. Still, knowledge regarding infestation control is low, and indiscriminate use of insecticides products were reported against domestic infestations of bed bugs, triatomines, and other synanthropic insects. Similar results were observed in peri-urban neighborhoods of the metropolitan Arequipa (40). Although householder application of insecticides was associated with lower risk of infestation by *T. infestans* in endemic area in Argentina (36), insecticide misuse might decrease susceptibility of vector populations to these and related products (41). Aerosol insecticide formulations, in particular, are easily available for purchase in Peru, and their use is widespread. These products often combine different insecticides and synergists agents and are unlikely to control triatomine infestations effectively. Use of aerosol products has shown to increase resistance to pyrethroids in populations of *Aedes aegypti* in Mexico (41). Health educational campaigns to improve community knowledge regarding methods to control synanthropic insects, particularly for bed bug infestations, could help to prevent misuse of insecticide products. However, until a safe and affordable alternative is made available for bed bugs, as it is for triatomines, it is hard to imagine the bed bug situation improving.

We suggest, based on our synthesis of spatial entomological data and our interviews, that the *Chirimacha* populations we detected and eliminated arrived at El Pedregal rather recently, and were hindered from dispersing by the numerous uninhabited lots in the growing city. It is possible that the insects are remnants of much earlier infestation processes that our participants recounted, but we believe that alternative explanation is less likely, because it is hard to imagine the insects being constrained to such small foci of infestations for such a long time. Indeed, in the neighboring district of La Joya, *Chrimachas* redispersed following control in the 1990s to infest over a quarter of households (42). It is also possible that the actions of residents applying their own insecticides prevented the further dissemination of *Chirimachas*, though again it is hard to imagine those activities would be effective and uniform enough for long term control. If we are correct our results are concerning—they suggest that as El Pedregal develops further, and the empty lots are filled in, the new city will be highly susceptible to triatomine infestations.

The historic and current triatomine infestations, which occurred about 30 years apart, seem to differ in their dispersal and density and the settings of space occupation. The former is recalled in interviews as a widespread event, affecting different neighborhoods, houses, and animal corrals at high densities. Housing conditions in those days were described as improvised constructions, using poor quality materials. The infestations we observed in Pedregal, however, were extremely spatially clustered, of low insect densities and only in a few houses that were constructed many years ago. Still, those houses were infested at low densities, and infestation spread was limited, likely by the environmental barriers created by unoccupied, or very sparsely occupied, lots, interspersed with more permanent and, for the insects, habitable, dwellings. Housing conditions, particularly construction materials and crowding, which provide availability of refuges and source of food for triatomines, are limiting factors for domestic infestation by triatomines, density of its colonies, and infection by *Try. cruzi* across many contexts (9,10,43–45). In the interval between the two (re-)introduction events, changes in housing materials and conditions in El Pedregal may not have made houses entirely refractory to infestation, but they hampered infestation, dispersal, and density of the insects.

Large government decisions involving water and changes in land use can have outsized impacts on the lives of farmers and other laborers, and these impacts can reverberate through society, leading to mass migration and, ultimately, increasing populations’ vulnerability to disease (46–49). The cost of these decisions is ultimately borne by those with the least recourse to bear them (50,51). Migration has been also driven by adverse climate conditions, and this phenomenon is expected to increase in the following years. It is urgent to advocate to a shift in governance that contemplates both a responsible use of common natural resources, and a preparedness to ensure food security to populations affected and displaced by climatic events.

Ironically for a city that grew around an irrigation project, many residents of El Pedregal lack access to clean water (16). Without running water many resort to storing water in barrels, which creates risk for the emergence of *Aedes aegypti* and Dengue virus (52). Emergence of vectors highlight the importance that communities and health authorities to remain hypervigilant, as the complex forces of migration and poor housing conditions makes El Pedregal especially vulnerable to vector-borne disease.

## Data Availability

All data produced in the present study are available upon reasonable request to the authors

## Acknowledgements

The authors gratefully acknowledge the contributions of the Ministerio de Salud del Perú (MINSA), the Dirección General de Salud de las Personas (DGSP), the Estrategia Sanitaria Nacional de Prevención y Control de Enfermedades Metaxénicas y Otras Transmitidas por Vectores (ESNPCEMOTVS), the Dirección General de Salud Ambiental (DIGESA), the Gobierno Regional de Arequipa, the Gerencia Regional de Salud de Arequipa (GRSA), the Pan American Health Organization (PAHO/OPS), the Canadian International Development Agency (CIDA). We also thank Amparo Toledo, Claudia Arevalo Nieto and Laura Tamayo for their valuable contributions to these studies.

